# ALICE: Improved speech in noise understanding with self-guided hearing care

**DOI:** 10.1101/2025.05.28.25328487

**Authors:** Astrid van Wieringen, Mira Van Wilderode, Les De Ridder, Tom Francart, Jan Wouters

## Abstract

**Objective:** Persons with hearing aids or cochlear implants often have difficulty understanding speech well despite amplification, especially in noisy environments. Auditory training can help their brain refine their listening skills. The current study aimed to determine the efficacy of the ALICE program, a self-guided home-based health care program including monitoring, training and counselling.

**Method:** A multicentric study was carried out, including hearing aid centers and a cochlear implant center in Flanders (Belgium). Participants were assigned randomly to an intervention or control group. Participants in the intervention group received a tailored flow of exercises that could be streamed to the device or presented in sound field. All participants were tested before and after 8 weeks using sentences in noise and different self-reports.

**Results:** Participants in the intervention group were compliant during the 8-week training period. Significant on-task improvements were observed, as well as improved speech in noise understanding for the intervention group only. The self-report data did not reveal changes following the intervention.

**Conclusions:** Our clinical trial shows that the self-guided ALICE training program is effective at improving the auditory system’s ability to parse untrained speech in noise. This enhancement in speech in noise performance is specific to the training group, as the control group did not improve. The results of the clinical trials imply that ALICE can be used as a scalable, accessible, and safe hearing care intervention.

## Introduction

According to the World Health Organization, over 1.5 billion people globally live with some degree of hearing impairment (HI), and around 430 million require rehabilitation services. This calls for efficient and cost-effective treatment (World Health Organization 2021). Without appropriate follow-up, a person with HI will experience increased listening effort and difficulties with communication, learning, social-emotional functioning, employment, and quality of life (Davis et al., 2016; Pichora-Fuller et al., 2016; Denham et al., 2024).

Regular care in hearing centers entails providing and fitting hearing aids as well as guidance on their maintenance. Auditory perceptual training is not offered unless at specialized audiological centers, while it is an essential cornerstone of hearing health care (Boothroyd, 2007, 2010). Auditory training is particularly important for people with HI and listening difficulties, as their brains may have diminished the ability to recognize the meanings associated with certain sounds. Persons with hearing aids or cochlear implants often have difficulty understanding speech well despite amplification, especially in noisy environments (Shehorn et al., 2018; van Wieringen et al., 2021). Auditory training can help their brain relearn how to process sound effectively. Additionally, when hearing sensitivity is good but the brain struggles to make sense of sounds, auditory training can help improve listening skills, spatial processing, binaural integration, and auditory memory (Schafer et al., 2024).

Counseling emotional and psychological challenges related to hearing loss and providing strategies for effective communication (Johnson et al., 2018) are also very important (Contrera et al., 2016; Timmer et al., 2024). However, time constraints, lack of knowledge, and limited reimbursement, among others, prevent implementing this in hearing health care for persons with hearing aids. The regular care for persons with cochlear implants is often different: most persons with a cochlear implant receive intensive rehabilitation during the first 6 months after their implantation, including auditory perceptual training and counseling in the clinic.

Over the past years, advancements in telehealth have led to accessible and tailored services for individuals with communication difficulties (Swanepoel & Hall, 2010; Swanepoel & Iii, 2020; van der Mescht et al., 2022). Developments in remote hearing health care accelerated during the COVID-19 pandemic. Self-guided home-based auditory training programs emerged as a practical solution for those seeking to practice and refine their listening skills at their own pace (Frisby et al., 2022; Sweetow & Sabes, 2007). Over the past years the efficacy of several programs was researched , such as LACE (Sweetow & Sabes, 2006, Saunders et al., 2016, Lai et al., 2023), Brain Fitness cognitive training (Anderson et al., 2013), ReadMyQuips (Abrams et al., 2015, Rishiq et al. 2016, Rao et al., 2017), a serious game scenario (Reynard et al., 2022), and LUISTER (Magits et al., 2023, Van Wilderode et al., 2023).

Despite a multitude of studies, no clear recommendations could be made yet on the functional benefit of auditory training, partly due to limited evidence and external validity (cf the systematic reviews Henshaw and Ferguson, 2013; Lawrence et al., 2018 for persons with HI and Dornhoffer et al., 2024 for CI users). As a result, many hearing healthcare professionals hesitate to promote auditory training. Many studies in these systematic reviews did not include a control condition or had a limited sample size. Auditory training aims to transfer skills to improve daily life communication and build confidence (Sweetow & Sabes, 2006; Ferguson & Henshaw, 2015), but the transfer to untrained (or real-life) conditions is very difficult to capture. This was also evident from a systematic review evaluating twenty-eight studies of self-guided (home-based) training programs with a control condition (van Wieringen and Van Wilderode, under review). Thirteen of the twenty-eight studies showed some transfer of learning to untrained materials. However, all studies reported improvements in the trained tasks, confirming that practice improves skills and abilities.

LUISTER was the first self-guided (home-based) hearing care program designed for the Dutch/Flemish language and operated on a tablet (Magits et al., 2023). It contained a monitoring module with the digits in noise (DiN) task and the vowel and consonant identification tasks (van Wieringen et al., 2021) and a training module with more than 500 Flemish/Dutch exercises. Bottom-up exercises focus on sound discrimination and identification, while top-down, synthetic, exercises were designed to develop higher-order skills using realistic materials such as sentences. The speech reception thresholds derived from the DiN task determined the starting level of the exercises for the individual, and the training exercises were tailored to address the client’s specific challenges. LUISTER was validated in adults with cochlear implants (Magits et al., 2023), as well as in individuals with hearing aids (Van Wilderode et al., 2023). The results of these studies provided scientific evidence supporting the added value of auditory perceptual training for people with HI. Notably, improvements to untrained measures were observed with the LUISTER training and an alternative program used by the active control group (Magits et al., 2023).

Building on this foundation, the monitoring and training modules were integrated into a comprehensive hearing care app called ALICE (Assistant for Listening and Communication Enhancement), supplemented with counseling and a dashboard for the hearing care professional (HCP). ALICE runs on smartphones (Android or iOS) and transmits data to a clinician’s dashboard via a web browser. Given the promising results of the LUISTER program, it is essential to evaluate the effectiveness of its successor, ALICE, in a real-world context. As a more comprehensive and accessible tool integrating monitoring, training, and counseling, ALICE can potentially enhance listening skills and empower persons with a HI. However, a randomized control trial is needed to assess its added value in routine audiological care. The training tasks and the tailored (personalized) flow chart were the same as those assessed with LUISTER (Magits et al., 2023; Van Wilderode et al., 2023). Unlike previous clinical trials, which were conducted at a single site, the present study was carried out across various hearing aid centers distributed throughout Flanders (Belgium), alongside the university hospital of Leuven. Furthermore, this study uniquely included an evaluation of the counseling module. Participants had a hearing aid or a cochlear implant and received either regular care (control) or regular care supplemented with the ALICE program. We expected that the tailored auditory training program would be engaging and promote adherence, and that it would enhance daily life listening and/or communication strategies by showing better speech in noise performance and reduced self-perceived listening effort. We also expected that the intervention group would be more knowledgeable about their HI and recognize the emotional consequences of their HI than the group without an intervention through the counseling module.

## Materials and methods

A priori power analysis was conducted using G*Power (Erdfelder et al., 2009) based on a repeated-measures ANOVA (α = 0.05). Given that G*Power does not support power calculations for linear mixed-effects models, this approach served as a reasonable estimate. It indicated that a total sample of at least 76 persons is needed to detect differences in speech understanding in noise between the two treatment groups with a low to medium effect size (d = 0.25). The data were subsequently analyzed using linear mixed-effects modeling to account for intrasubject variability, providing a more accurate representation of the experimental design.

### Participants

The hearing care professional (HCP) at the local investigation site recruited adult clients with at least 6 months of hearing aid experience, and the audiologist at the university hospital recruited CI users with at least 6 months of experience. All participants were Dutch-speaking and were able to use the smartphone or tablet, and had sufficient eyesight to read the screen.

### Procedure

Randomization and study data were managed using Research electronic data capture (REDCap) hosted at KU Leuven, the main investigator. REDCap is a secure, web-based software platform to support data capture for research studies (Harris et al., 2009; 2019). Two sessions were planned, one before and one after 8 weeks. After consent to participate in the study, the speech threshold for sentences in noise was determined twice, and the questionnaires were filled out by scanning a QR code and responding to the questions online (session 1). Subsequently, the local investigator (HCP) would send the age and gender of the participant to the main investigator for randomization in REDCap. Subsequently, participants in the intervention group were shown how to download and use the ALICE program. All participants received a new appointment for sentence understanding in noise and filling out self-report after 8 weeks (session 2). Participants in the control group were given the chance to do the training tasks afterwards.

Ethical approval was granted by the Federal Agency for Medicines and Health Products (Eudamed number: CIV-22-06-039801-SM03). Participants were not paid. The study protocol is registered on ClinicalTrials.gov (NCT05329922). This study complies with the Consolidated Standards of Reporting Trials extension for nonpharmacologic treatments (Boutron et al. 2017).

### Demographics

Initially, 68 participants were included in the ALICE intervention group. Three participants wished to stop (1 person due to persistent technical difficulties, 1 person due to the difficulty of the app increasing too quickly, and 1 person due to persistent tinnitus before the start of the clinical trial). Sixty-nine persons were assigned to the control group. Four people decided to stop participation (3 due to adverse events (an ear infection, reimplantation of CI, and fungal infection so that the participant could not wear hearing aids for a while), one without reason.

The demographics of the participants who completed the clinical trial are listed in Table 1. Each group contained 65 participants. CI users had more than 6 months of experience with their device and had completed their clinical rehabilitation. All participants presented with a postlingually acquired profound HI, and they communicated through spoken language in their daily lives. The number of CI and HA users was evenly distributed over the two groups. The two treatment groups did not differ in terms of chronological age, gender, type of hearing device, and other relevant factors (Table 1). Most participants were not professionally active anymore. Speech materials could be streamed to the device or presented in sound field. Participants with bilateral devices chose the side where they wished to stream the speech materials.

**Table I.** Demographics of the participants in the two groups (ALICE and Control). The table lists their hearing device (CI=cochlear implant, HA= hearing aid), gender, age distribution, working or not, severity of unaided hearing loss (HL, mild = pure tone average (PTA) < 40 dB HL, moderate: PTA between 40 dB HL and 70 dB HL, and severe: PTA > 70 dB HL).

|  | **ALICE** | **Control** |
| --- | --- | --- |
| **TOTAL** | 65 | 65 |
| **Hearing device** |  |  |
| CI and HA | 8 | 9 |
| Bilateral HA | 50 | 50 |
| Unilateral CI | 1 | 1 |
| Unilateral HA | 6 | 5 |
| **Gender** |  |  |
| Female | 31 | 31 |
| Male | 34 | 34 |
| **Age group** |  |  |
| 18-40 | 4 | 3 |
| 41-60 | 14 | 14 |
| 61-80 | 43 | 45 |
| **Severity of HL** |  |  |
| Mild HL | 39 | 23 |
| Moderate HL | 15 | 29 |
| Severe HL | 7 | 10 |
| Not professionally active | 46 | 40 |
| Professionally active | 19 | 25 |
| Android | 39 | 42 |
| IOS | 26 | 23 |
| Streaming | 37 |  |
| Sound field | 28 |  |

### The ALICE program

The ALICE program contains two parts: 1) a client application that can be downloaded onto a personal device (smartphone or tablet), and 2) a web-based dashboard for the HCP with data and analytics of their clients. The client application comprises three modules: a performance monitoring module, a listening training module, and a counseling module. The monitoring tasks are the digits in noise (DiN) task and the phoneme discrimination task described in Magits et al. (2023). The speech reception threshold (SRT) of the DiN task is used to determine the starting level or signal-to-noise ratio of a training task. The DiN task was administered at the beginning of each week, followed by either the vowel or the consonant identification task in quiet (without feedback). This task’s most difficult discriminable pairs were assigned a greater weight in the training tasks. The results of both tasks are displayed on the HCP’s dashboard.

The *listening training exercises* are divided into five categories: vowels, consonants, themes (e.g., bathroom, kitchen, …), suprasegmental (including voice recognition and emphasis), and sentences (clock reading, identifying similar sounding words in sentences, realistic sentences). Exercises are performed in quiet and in various types and levels of background noise in a closed-response set (3, -6 or 9 alternatives). They are varied so that participants do not perform the same exercise for more than 4-5 minutes. Feedback is provided after each item, and the speech sound can be replayed if desired to refine the listening skills. A predefined set of rules determines whether the participant needs to repeat a similar task or is referred to an easier or more challenging one (Magits et al., 2023). Participants received feedback after each trial but were not shown absolute scores. Task progress was tracked via the dashboard. Participants started training in quiet, and when they performed sufficiently on these tasks, similar tasks were presented in noise, specifically speech-weighted noise (SWN), then babble noise (BN). This approach resulted in a tailored selection of tasks and difficulty levels for each participant.

With the *counseling module* the participant is requested to respond to different questions about his/her listening experience and difficulties. These questions are based on validated questionnaires such as the Speech Spatial and Quality of Hearing questionnaire (Gatehouse and Noble, 2004), the Hearing Handicap Inventory for Adults/the Elderly (Ventry and Weinstein, 1982), and the Fatigue Assessment Scale (De Vries et al., 2004).

At the beginning of the training program, the participants in the intervention group indicated difficult listening conditions from a closed set of responses (difficulties at home, at work, during cultural activities, group conversations, leisure, music, one-on-one conversations, outdoors, sports, telephone, and/or television). Subsequently, up to three questions linked to these questions were presented daily to assess their listening experience. Questions were answered on a 5-point Likert scale. Responses were categorized according to ‘ability’ (‘I can follow a conversation during dinner’), ‘feeling’ (’Because of my hearing loss, I feel stressed’), and ‘participation’ (‘I avoid groups because of my hearing loss’). Results are sent to the HCP dashboard.

### Outcomes used in the randomized clinical trial

The primary outcome was a validated sentence understanding in noise measure. This main outcome was complemented with questionnaires to evaluate effectiveness in a broader context of functioning and participation.

### Sentence understanding in noise

Sentence understanding in noise was assessed with the LIST sentences (van Wieringen & Wouters 2008). Sentences were presented in speech-weighted noise: the noise level was kept constant at 65 dB SPL. Starting at 55 dB SPL, the level of the first sentence was increased in steps of 2 dB until the sentence was identified correctly. Subsequently, the intensity level varied adaptively during the following nine sentences with a one-down-one-up procedure to target the 50% point on the psychometric function (speech reception threshold, SRT). The SRT is calculated based on the last five responses plus the imaginary last SNR level. The test was presented twice to the listener, both before and after the 8-week period, to control for procedural effects. Sentences were presented in sound field via a loudspeaker. Lists of sentences are counterbalanced and never presented twice.

### Questionnaires

The SSQ12 is a short version of the validated Speech, Spatial, and Qualities of Hearing Scale (SSQ, Noble et al., 2013). The SSQ assesses a person’s hearing ability in everyday life, particularly in complex listening environments. In the 12-item questionnaire, the first five questions belong to the subscale ‘speech’ (speech in noise, multiple speech streams, speech in speech). Items 6,7,8 capture self-perceived benefit on localization, distance, and movement, and the last four questions refer to segregation, identification of sound (musical instrument), quality, and listening effort. The higher the score, the better the self-perceived performance. Participants scored each question on a visual analogue scale of 0 to 10. We expected that these domains would be rated higher after the training intervention.

The CAS (Communication and Acceptance Scale) is a validated scale comprising 18 items (Öberg et al., 2021). It was developed to detect clinical changes in “communication strategies and the emotional consequences, knowledge and acceptance of HI”. ). It contains 18 items on a 5-point scale: completely agree (4), agree (3), neutral (2), disagree (1), and completely disagree (0). Responses are cast into four subscales: emotional consequences, verbal communication strategies, confirmation strategies, and hearing knowledge and acceptance. Higher scores indicated better functioning (Öberg et al., 2021). The statements are divided into 5 subscales: confirmation strategies (Conf, 3 questions), Emotional consequences (Emo, 8 questions), Hearing Knowledge (HKnow, 1 question), hearing loss and acceptance (HAccep, 4 questions), and Verbal communication strategies (VerbalC, 2 questions). This questionnaire has been translated from Swedish to Dutch (and back-translated to ensure correct translation. It was anticipated that the more knowledge (subscale 4) participants obtained through counseling and learning communication strategies, the better prepared they would be to use the strategies in real life (subscales 2 and 3).

EAS (Effort Assessment Scale) is a validated self-report questionnaire on listening effort (Alhanbali et al., 2017). It captures how much mental energy and cognitive effort are required for a listening task. Six questions on different listening situations are presented. Participants rate their effort on a visual analogue scale, from 0 (no effort) to 10 (extreme effort). The EAS questions were translated into Dutch for this study.

The IOI-HA (International Outcome Inventory for Hearing Aids) assesses the daily use of hearing aids (or cochlear implants), as well as the perceived benefit and satisfaction. The IOI-HA consists of 7 questions rated on a 5-point Likert scale and has been validated in Dutch (Kramer et al., 2002). Each question addresses a different aspect of hearing aid use (daily use, benefit in different listening situations, activity limitation, satisfaction, participation restrictions, impact on relationships, and quality of life). The higher the value, the more positive the respondent is about a situation.

### Data Analysis

Statistical analyses were performed using the R programming language and statistical environment (R Core Team 2024). Potential improvements between the first and second sessions were assessed using linear mixed models (LMMs). The participant was considered a random factor, and session and treatment group (intervention versus control) were fixed factors.

## Results

The current study aimed to determine the efficacy of the ALICE program. Below, we present the results for the CI users and HA users separately. Progress on exercises, the DiN task and vowel and consonant identification was derived from the HCP dashboard. The clinical trial outcomes were documented in REDCap.

### Adherence and exercises

Fig. 1 illustrates the total practice time per participant for the 9 CI participants (left) and the 56 HA participants (right). The black horizontal line indicates the recommended total practice time during the 8 weeks. Most participants adhered to their schedule (or practiced more than recommended), and about 5 participants practiced less than half of the recommended time.

**Fig. 1.**
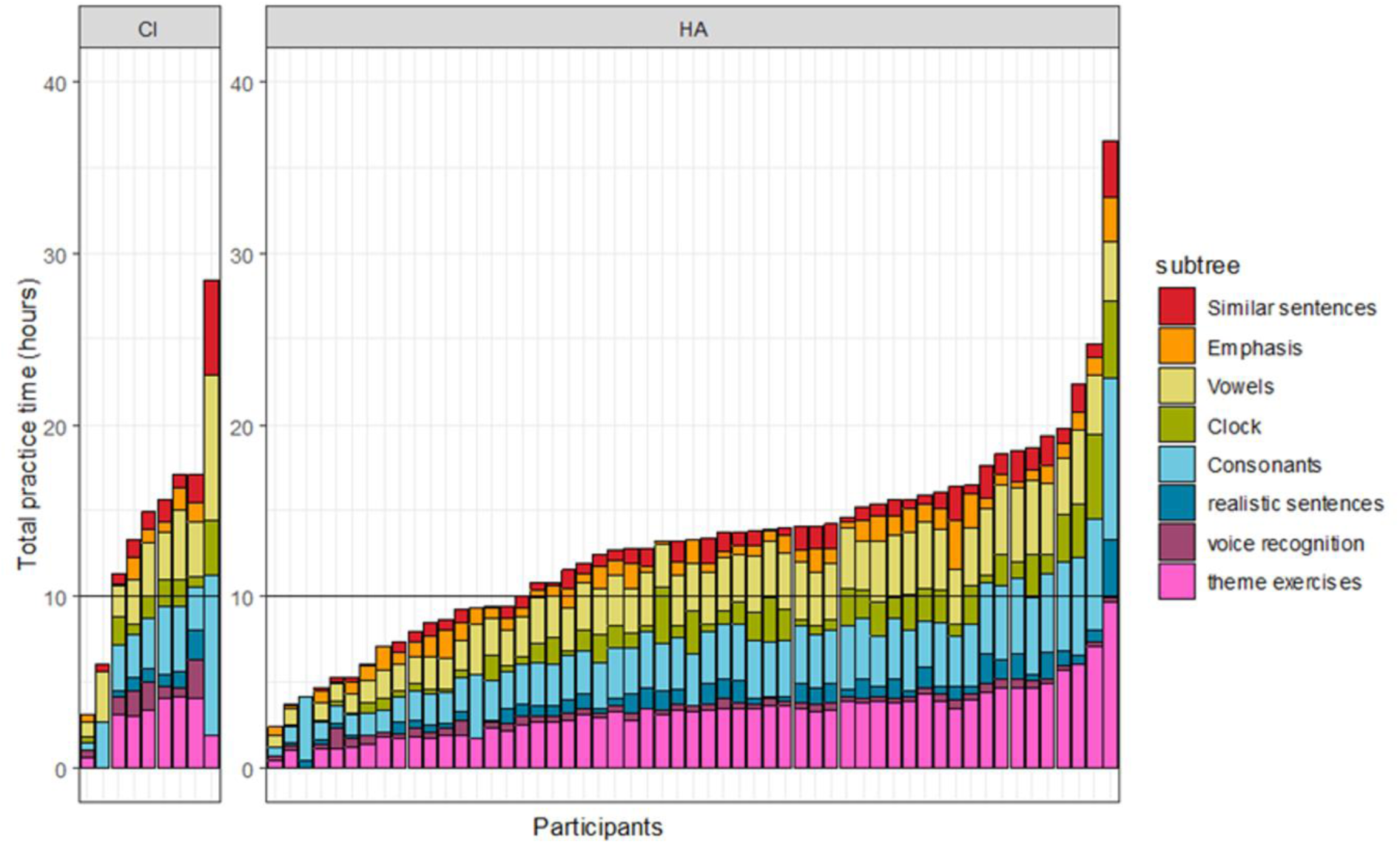
about here. Adherence to training and time spent on exercises for CI (=9) and HA (n-57) users. The black horizontal line indicates the recommended total practice time during the 8 weeks

### On-task improvements for the ALICE group

At the beginning of a training week, participants first performed a digits-in-noise task and alternating, either a vowel or consonant identification task. These data were logged in the HCP dashboard. Fig. 2A illustrates the SRTs of the CI and HA users for the eight weeks of training. Error bars depict two times the standard error between participants. As expected, the average SRTs of the CI users were higher than those of the HA users, although some CI participants performed in line with HA users. However, both groups improved with time (β=-0.32 SE = 0.11, t= -2.8, p =0.005).

**Fig 2a.**
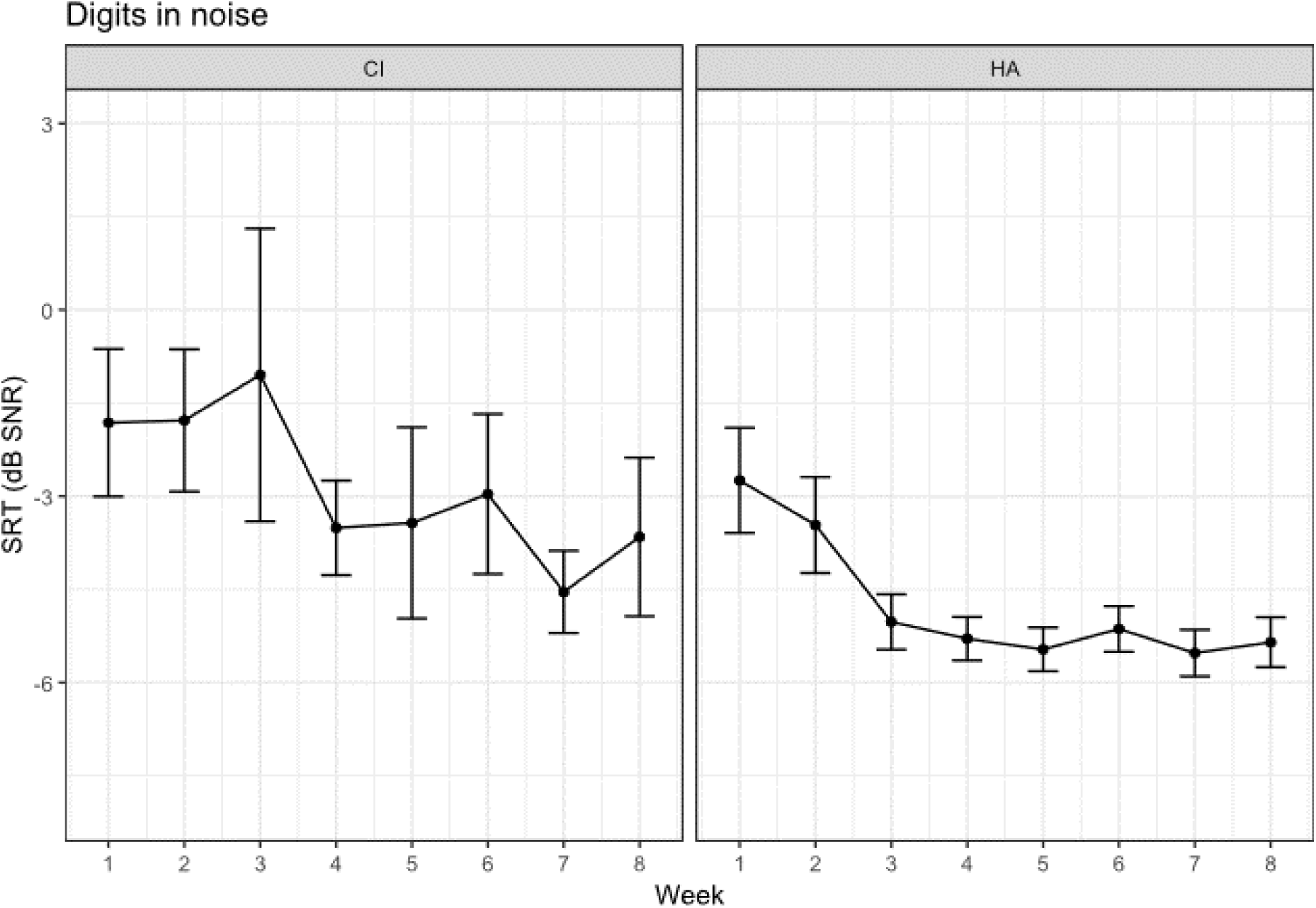
Average SRT (and standard error) with the DiN task during the eight weeks of training for the CI (n=9) and HA (n=56) users. The number of participants in the final weeks is slightly lower due to diminished adherence. **

Fig. 2b shows the percentage correct scores for vowel and consonant identification tasks for CI and HA participants over 4 periods (8 weeks). Despite high scores, they are not perfect, indicating room for improvement. CI users scored lower than HA users for both vowel (β= 16.2, SE = 3.1, t= 5.2, p <0.001) and consonant identification (β= 19.4, SE = 5.6, t= 3.5, p <0.001). Both groups improved over time for vowel (β= 0.63, SE = 0.29, t= 2.2, p =0.03) and consonant identification (β= 1.3, SE = 0.32, t= 3.8, p <0.001).

**Fig. 2b.**
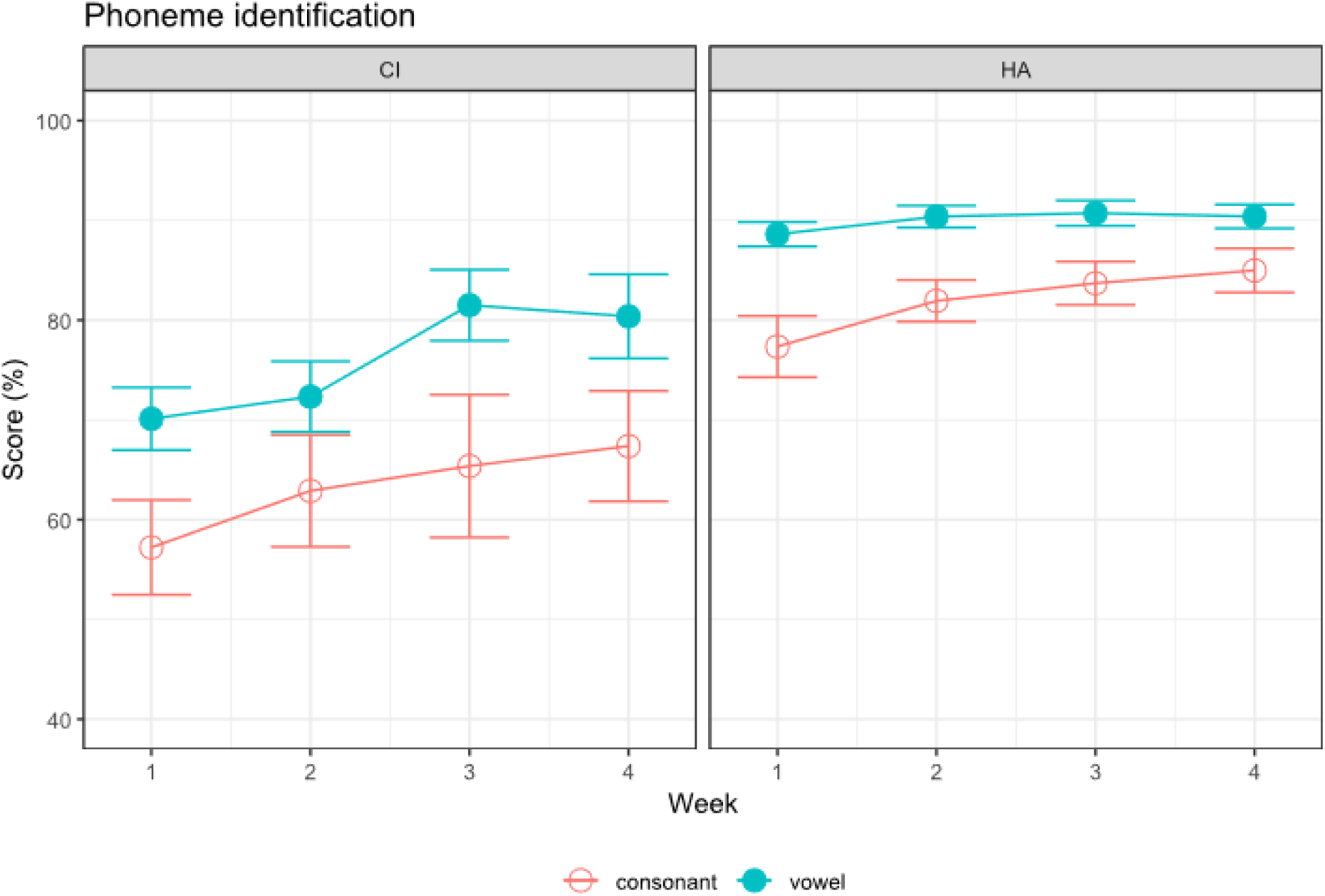
Percentage correct phoneme identification for CI (n=9) and HA (n=56) participants for 4 periods (8 weeks) during the intervention. The number of participants in the final weeks is slightly lower due to diminished adherence.

### Transfer to untrained materials

#### Speech-in-noise understanding

An important question is whether the training gains transfer to functional benefits in real-world listening and whether these improvements are only present in the intervention group. These functional benefits can be improvements in (untrained) speech in noise understanding, and memory, attention, and self-perceived benefits. Fig 3 illustrates the performance on speech understanding in noise before and after auditory training for each participant separately. The area between the full lines represents changes in speech understanding in noise that fall within -2 and +2 dB SNR, indicating a clinically relevant improvement or decrement.

**Fig. 3.**
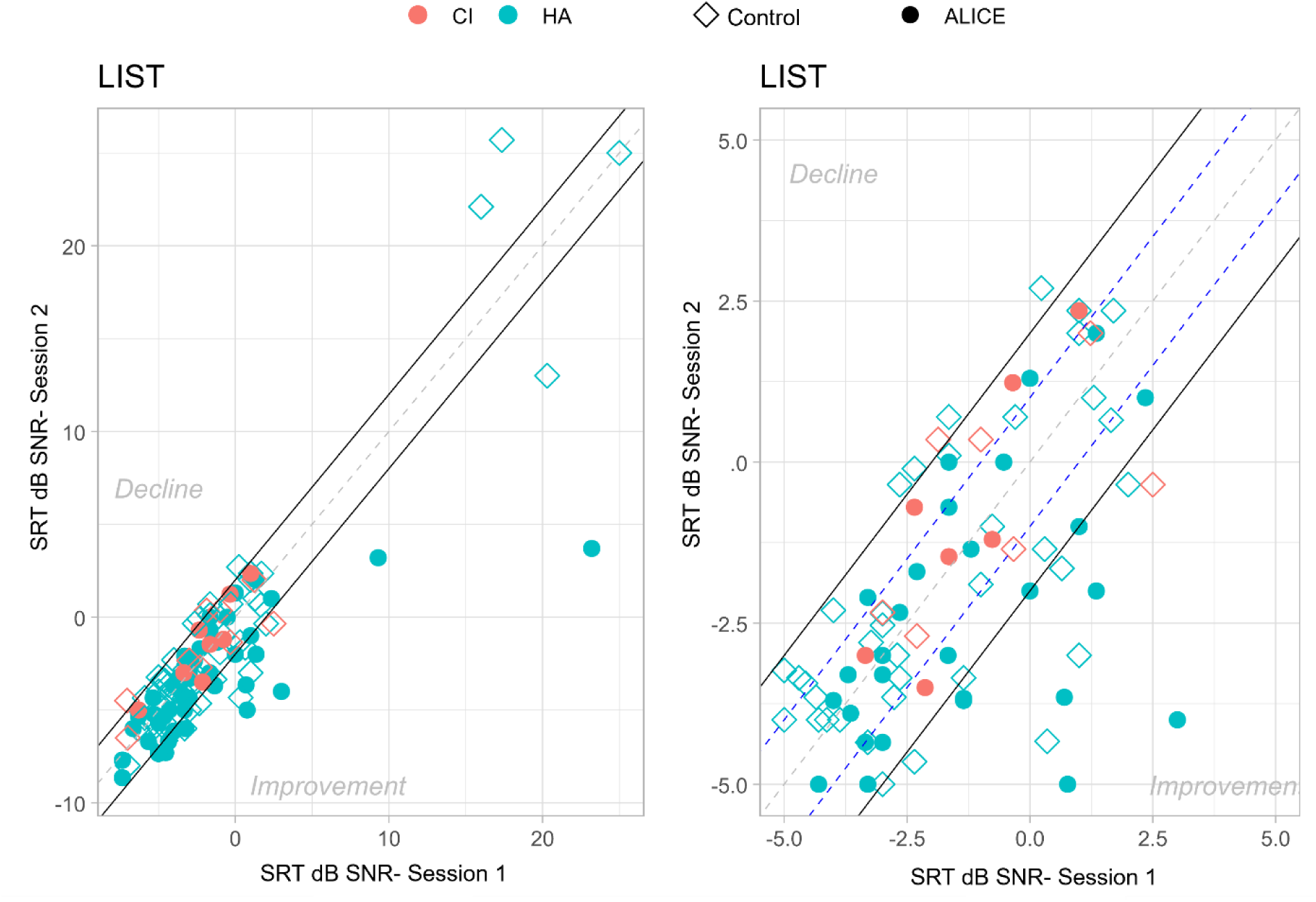
speech understanding in noise before (session 1) and after (session 2) auditory training for each participant separately. The area between the full lines represents changes in speech understanding in noise that fall within a range of -2 to +2 dB SNR. The plot on the right shows a zoomed-in portion of the plot on the left, without the outliers in the top right corner.

An LMM analysis on the SRTs of the LIST sentences, with treatment group and session as fixed factors and participant as random factor, yielded a significant effect of treatment (β=-1.98 SE = 0.93, t= -2.12, p =0.03) and a significant effect of session (β=-0.51, SE = 0.19, t=2.67, p =0.008). Moreover, the two-way interaction of session and treatment was significant (β=-1.12, SE = 0.38, t=-2.92, p =0.004). Post-hoc tests following the two-way interaction with Bonferroni corrections showed that session 1 was not statistically different for the two treatments (ΔM=1.31 dB SNR, t(121)=1.36, p=0.17) and that the difference in sentence understanding in noise between the control group and the ALICE intervention was significant after training (ΔM=2.51 dB SNR, t(115)=2.50, p=0.01).

Further analysis revealed no significant link between time spent on training and speech understanding in noise improvements (Spearman ρ=0.03, p=.89). However, individuals with better unaided PTA improved more than those with worse PTA in the intervention group. This correlation remained significant even after adjusting for age (partial correlation): r=0.32, p=0.02, F(53)=2.41 for the ALICE training group.

#### Self-Perceived benefit

Questionnaires were administered to assess whether use of the ALICE program would lead to improved self-perceived communication skills and quality of life. Below, we report the data from the four questionnaires. The number of responses per questionnaire may vary. To alleviate the workload of HCPs, participants could fill out the questionnaire by scanning a QR code. However, this method also resulted in some missing values. Only fully completed questionnaires from the same participant for both session 1 and session 2 were taken into account. Each figure indicates the number of responses per questionnaire.

Fig 4. Illustrates the average performance scores (and standard errors) for session 1 and session 2 for the intervention and the control group. A LMM analysis on the performance scores with treatment group and session as fixed factors and participant as a random factor showed significant differences between different questions, because they tap into different domains. For example, SSQ3 (‘speech’) is significantly different from SSQ11 (‘quality’) in both the ALICE intervention group and the control group). However, neither the two groups (treatment vs control), nor the sessions, were significantly different. There were no significant interaction effects.

**Fig. 4.**
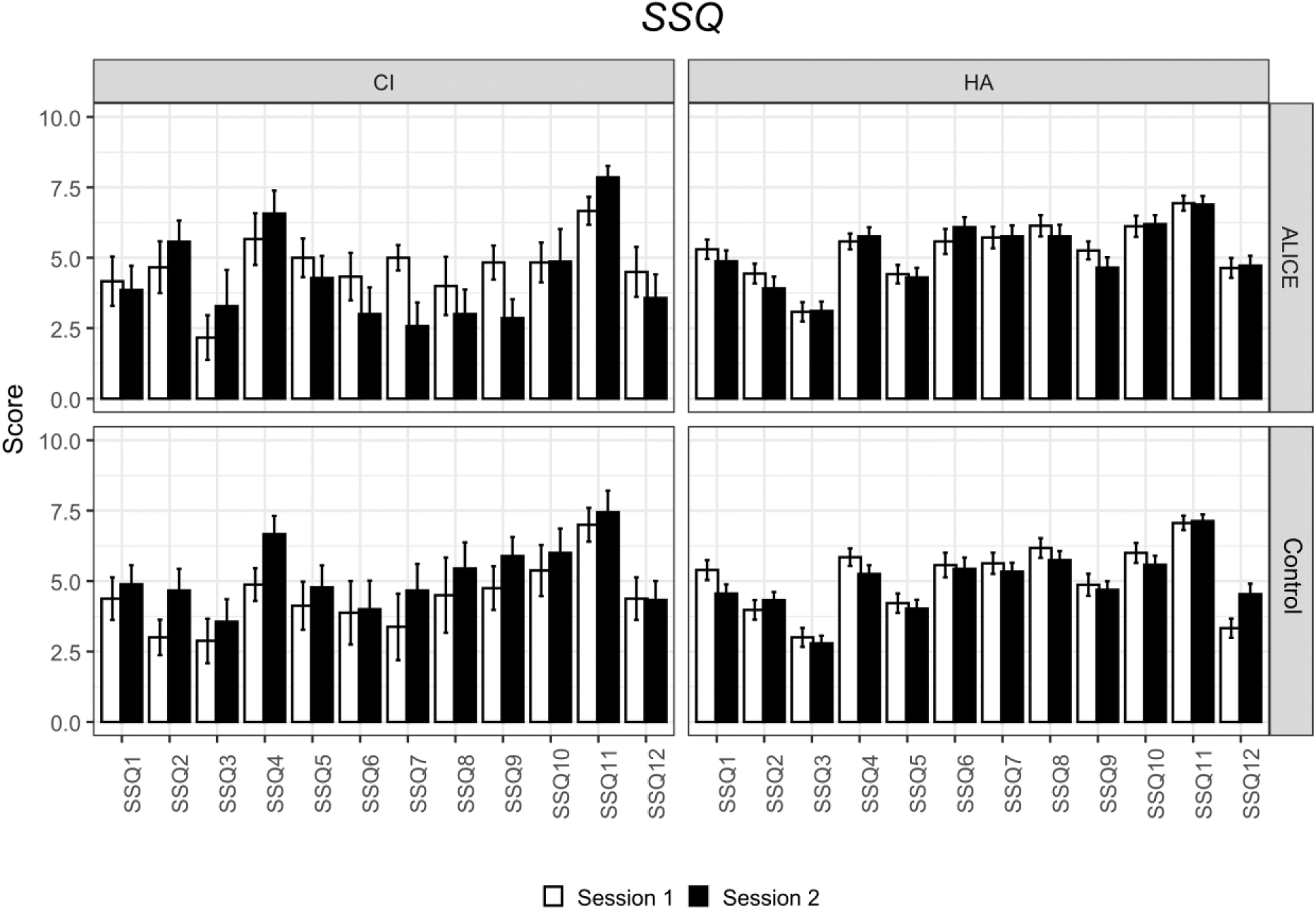
Average response scores for each question are shown for the intervention and control groups: Session 1 – CI (n = 8), HA (n = 56); Session 2 – CI (n = 8), HA (n = 56).

Although this questionnaire does not show improvements after training, it is noteworthy that the scores are relatively low, indicating that individuals are aware of their listening difficulties. This is especially the case for question 3 (when you have a conversation with one person in a room where there are many other people talking. Can you follow what the person you are talking to is saying?).

Fig. 1 also illustrates the amount of time spent per exercise type. Most time was spent on the theme exercises (e.g., kitchen utensils, bathroom, animals), but also on minimal pairs (consonants and vowels), followed by suprasegmental tasks (voice recognition and emphasis), and sentences. The figure does not depict the task sequence.

#### Communication and acceptance scale

The communication and acceptance scale (CAS) is intended to capture changes in communication strategies and the emotional consequences, knowledge, and acceptance of hearing loss.

A LMM analysis of the performance scores with treatment group and session as fixed factors and participant as random factors showed significant differences between different subscales, but not between groups (ALICE vs control) or between sessions (pre versus post). There were no significant interactions.

It’s quite notable that the knowledge question (Hknow) “I feel like I have a good understanding of my hearing loss” received a low rating (< 5 of a (re)scale of 10) despite counseling and practical advice. This may indicate that limited knowledge affects the ratings of verbal communication strategies and acceptance. Confirmation (Conf) pertains to two items representing verbal and psychosocial acknowledgment of hearing loss. Interestingly, only the subscale for emotional aspects of hearing loss was rated relatively higher, perhaps because they were volunteers keen to participate in the study.

#### Effort Assessment Scale

The EAS consists of 6 items, each targeting different listening situations (in quiet, in noise, during conversation). Higher scores indicate greater perceived listening effort. We anticipated that training would reduce listening effort. Our results do not support this hypothesis, as illustrated in Fig 6. Ratings are similar for session 1 and session 2 for both groups. Participants in both groups indicated high listening effort on most questions, especially question 4 ( Do you have to put in a lot of effort to follow discussion in a class, a meeting, or a lecture?) and question 5 (Do you have to put in a lot of effort to follow the conversation in a noisy environment (e.g., in a restaurant, at family gatherings)? Listening effort in conversation with others (question 1), amount of concentration when listening (question 2), ability to ignore other sounds (question 3), and listening on the telephone (question 6) appear somewhat less effortful.

**Fig. 5:**
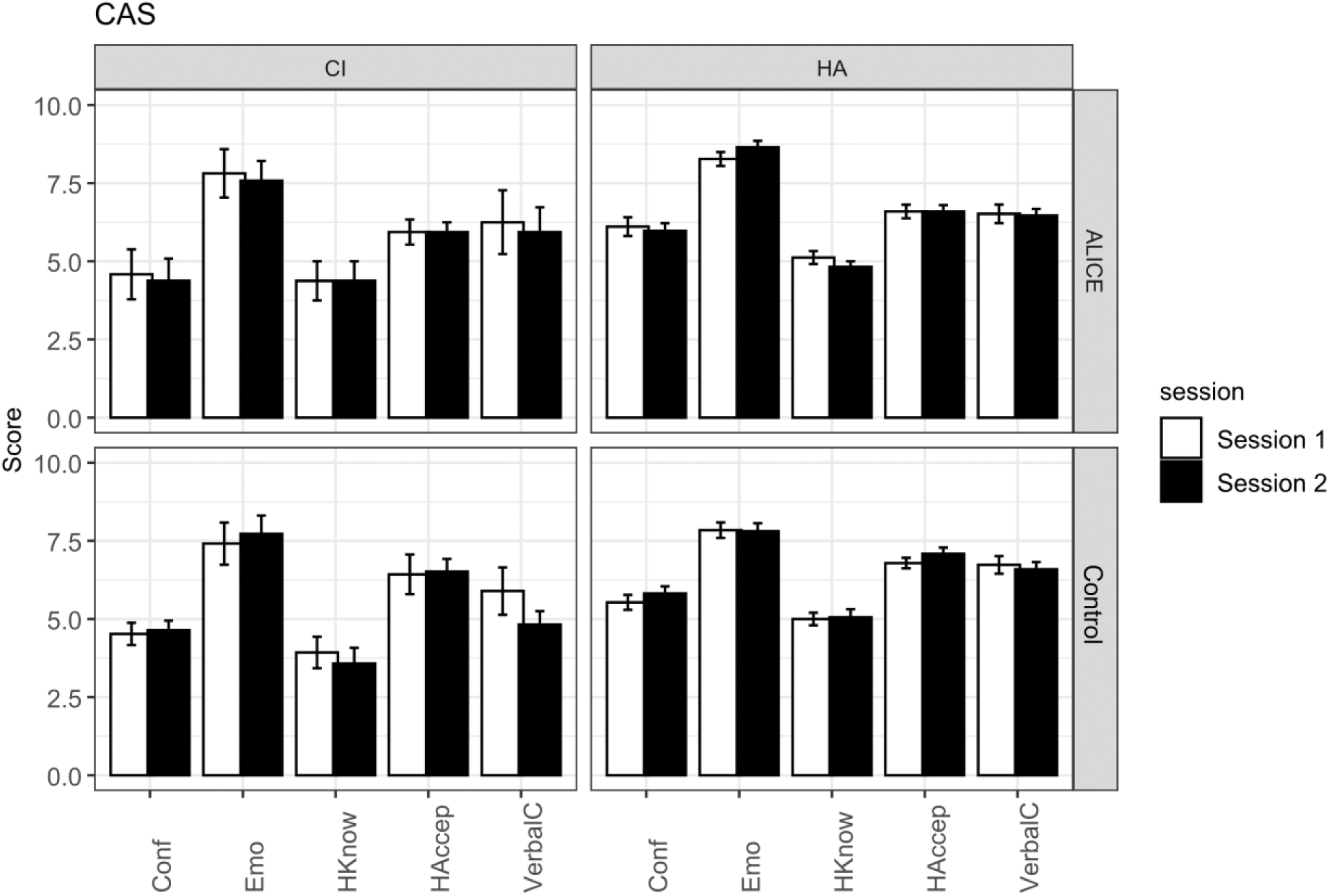
Average performance (and standard error) for each of the 5 subscales for the intervention and control groups: Session 1 – CI (n = 4), HA (n = 42); Session 2 – CI (n = 7), HA (n = 44). Scores of the 5-point likert scale are rescaled from 0-10.

**Fig. 6.**
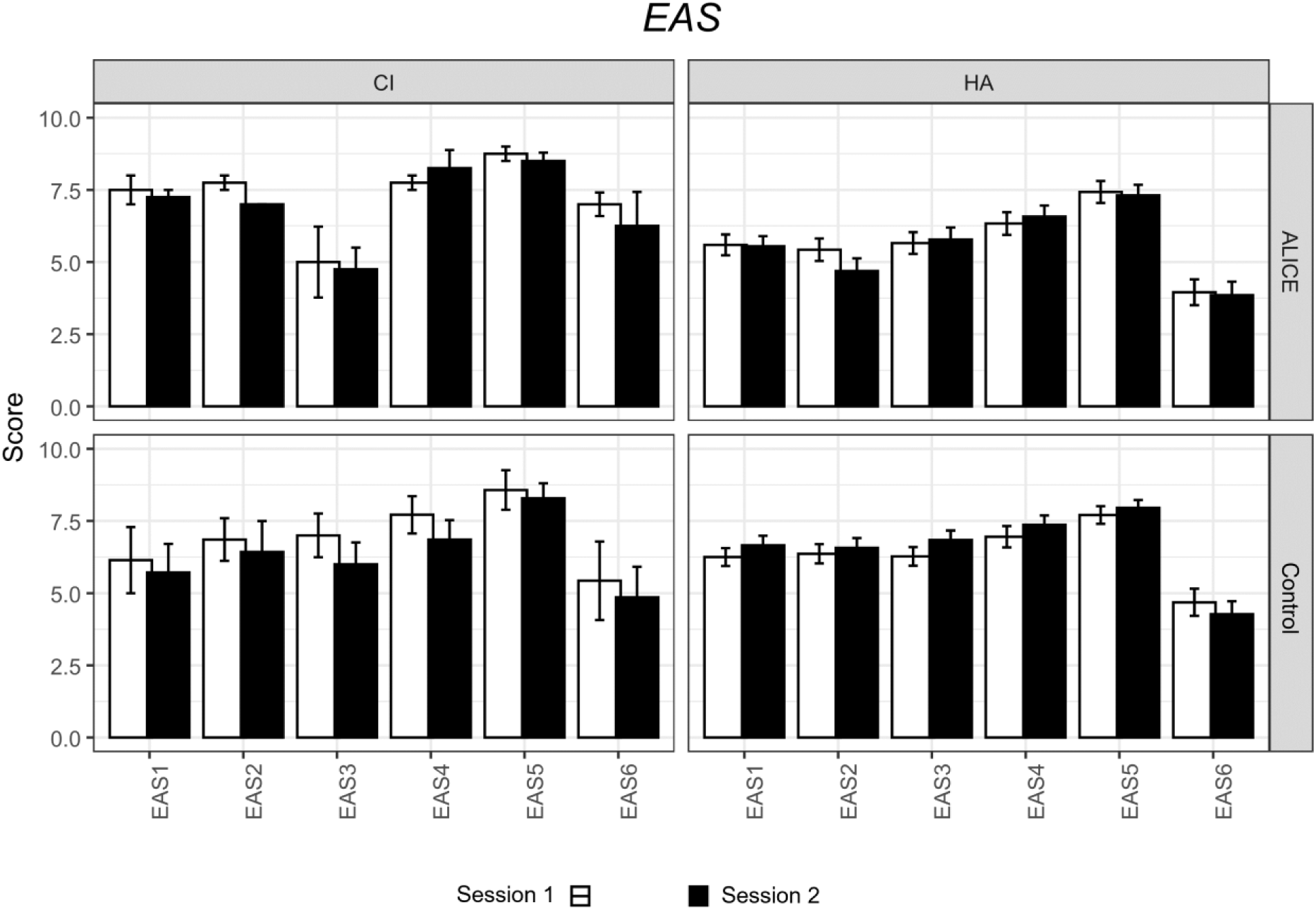
Average performance (and standard error) on the scores of the EAS for the intervention and control groups: Session 1 – CI (n = 4), HA (n = 44); Session 2 – CI (n = 7), HA (n = 42).

#### International Outcome Inventory for Hearing Aids

We expected that some perspectives on hearing aid use may change after the intervention. However, the LMM analyses showed significant differences between questions, but no significant main effect of group or session (Fig. 7).

**Fig. 7.**
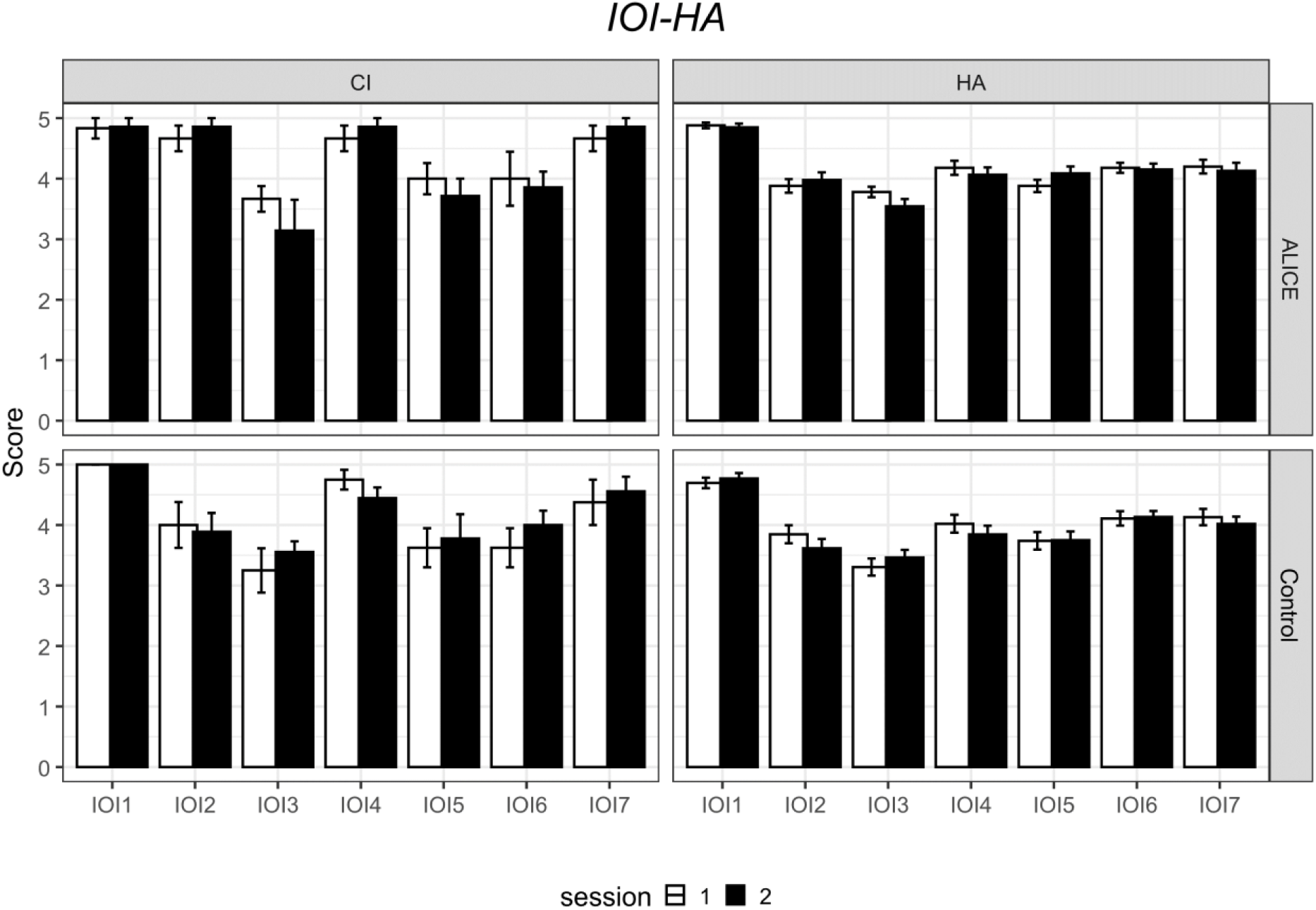
Average performance scores (and standard error) on the 6 questions of the IOI-HA for the intervention and control groups: Session 1 – CI (n = 7), HA (n = 51); Session 2 – CI (n = 4), HA (n = 50)

#### *Counseling* (only for the ALICE group)

Before the training with ALICE, participants indicated where and which activities they encountered difficulties with. Table II lists a summary of the difficulties noted by participants at the start (n=65). Most participants indicated encountering difficulties at home (note that only a few participants were professionally active) and mentioned struggling with conversations (both one-on-one and in groups) and watching television. Subsequently, participants received questions on these topics throughout the training period. These questions referred to their ability, feelings, or participation.

**Table II.** Counselling locations and number of participants who indicated having problems (Number)

| Location | Number of participants encountering difficulties |
| --- | --- |
| At home | 51 |
| At work | 15 |
| *Note: only a few participants are still working* |  |
| Culture | 25 |
| Group conversations | 57 |
| Leisure | 45 |
| Music | 38 |
| One-on-one-conversations | 58 |
| Outdoors | 51 |
| Sports | 21 |
| Telephone | 47 |
| Television | 54 |

During the clinical trial, participants received up to three questions per day related to either ’participation,’ ’ability,’ or ’feeling.’ Figure 8 shows the average ratings for these activities across the three categories during the 8 weeks of the trial. Higher scores indicate better self-reported values. Ratings did not differ significantly for the 3 different categories, nor as a function of time.

**Fig. 8.**
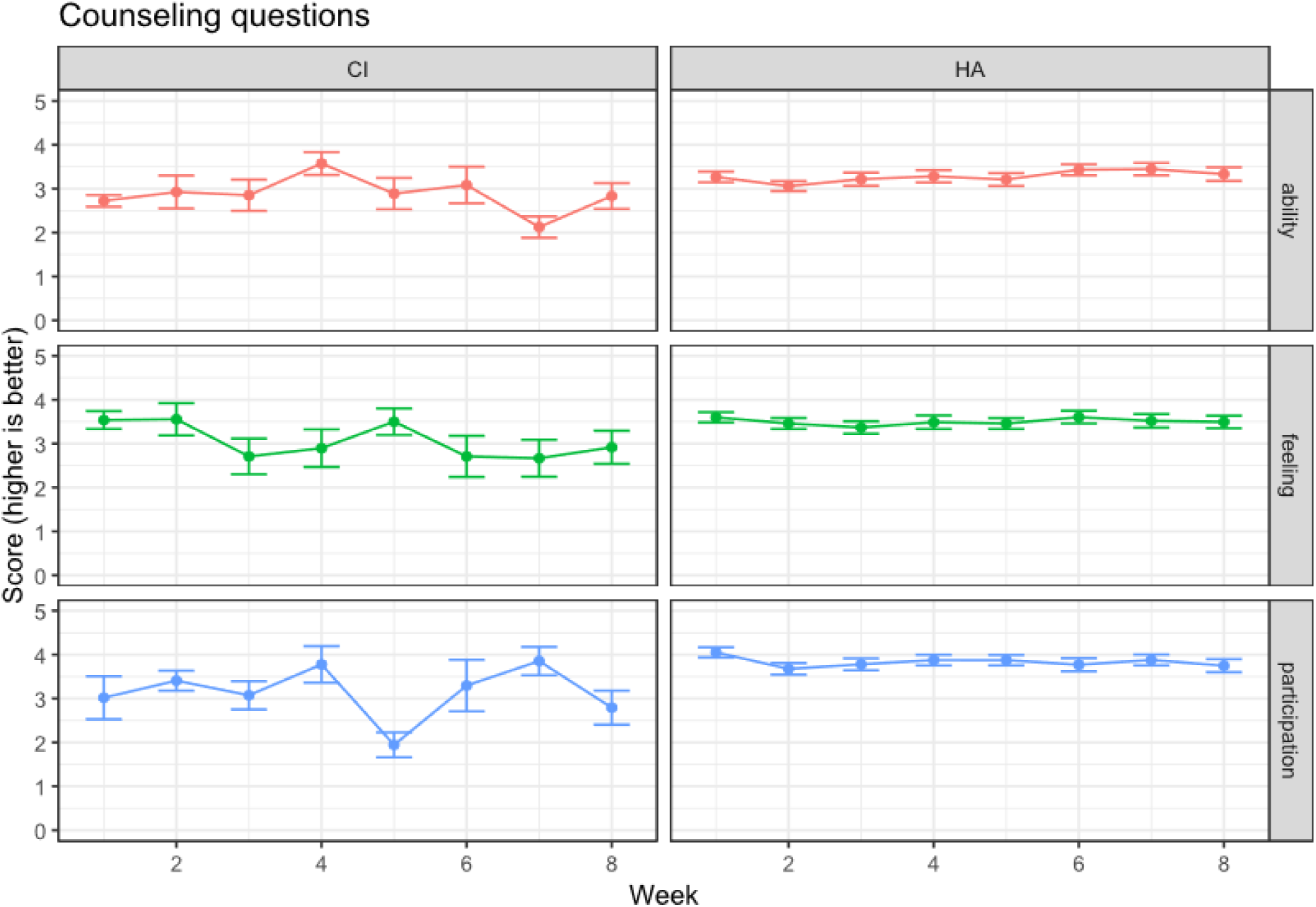
Average ratings (and standard error) on counselling questions for the CI and HA users during the 8-week intervention period. Counselling questions reflected on ‘ability’, ‘feeling’, and ‘participation’.

## Discussion

Auditory training programs are designed to improve a person’s ability to understand and interpret (speech) sounds. They are designed on the premise that attention-driven stimulation adapts the listening brain. Due to multiple exposures to sounds or speech at a tailored level or signal-to-noise ratio, sound patterns are reinforced. With practice, the brain forms or strengthens neural pathways to detect subtle differences in sound better, improve speech recognition (especially in noisy environments), and enhance auditory memory and attention (Russo et al. 2005; Anderson and Kraus, 2013; Ying et al. 2025).

Our clinical trial shows that the self-guided, tailored ALICE training program enhances DiN and phoneme perception, indicating that auditory training strengthens sensory processing. The training program is also effective at improving the auditory system’s ability to parse untrained speech in noisy conditions—a common and challenging real-world problem for individuals with HI. This enhancement in speech in noise performance is specific to the training group, as the control group did not improve. However, the ratings on the counseling questions (ability, feeling, participation) did not change with time. Contrary to our expectations, no reduction in self-perceived listening effort was observed, nor improvements in self-perceived benefits for the intervention group. The intervention group did not report an increased knowledge about their hearing impairment and did not recognize the emotional consequences of their HI more after using ALICE than the control group. Although improvements were noted in speech tasks, participants as a group did not report feeling less effort or experiencing greater benefits in their daily lives.

The discrepancy between behavioral performance gains and subjective experiences is well-known (Cox & Alexander, 1999; Sweetow Sabes, 2006). Questionnaires provide insight into how listening difficulties affect the quality of life and confidence, but they can also be influenced by personality, situational factors, and expectations (Patry, 2011). Moreover, it could be that participants are not aware of the beneficial changes that occur.

Bridging the disparity between behavioral gains and subjective experiences is crucial in hearing health care. To increase the likelihood of self-reported benefits, real-life communication simulations could be included in the training (e.g., conversations in cafés, group chats), or participants could customize listening scenarios based on their everyday challenges. If the training feels more like real-life listening, improvements may transfer more directly to daily communication. Additionally, functional benefits could be evaluated using virtual reality to mimic real-world listening scenarios for a more nuanced assessment (e.g., Devesse et al., 2020).

Another option would be to teach participants how to notice and use their improvements (e.g., communication strategies provided by the counseling module) by including reflective components. People may not perceive improvements if they do not recognize when and how they’re doing better. Integrating self-monitoring and reflection into auditory training can influence outcomes based on how well listeners are trained to recognize and adapt to their performance (Imhof, 2001). Currently, counseling questions about participation, feelings, and abilities are not connected to performance on the ALICE exercises. Introducing regular feedback check-ins could be valuable. In the future, individuals might be encouraged to reflect on their abilities, emotions, and level of participation after completing an exercise, helping to increase self-awareness

Finally, more sensitive self-report measures are required. Many of the self-report measures are not sensitive to changes in performance and may not be the right ones to capture subjective improvements after training. Instead of using questionnaires to capture subjective experiences, it may be more effective to use ecological momentary assessment (EMA, Schinkel-Bielefeld et al., 2024). EMA captures subjective experience in the moment, avoiding recall by asking short questions throughout the day (“How hard was it to understand someone at work today?). EMA may capture small day-to-day shifts in perceived effort or confidence that traditional questionnaires might miss.

Our current trial data show that adherence is good with the self-guided ALICE training. However, variability in adherence and dropouts remains a concern, and more research is needed to understand the factors underlying motivation and engagement. Engagement and adherence to computer-based auditory training are influenced by intrinsic motivation (e.g., hearing difficulties) and extrinsic motivation (e.g., the desire to help others with hearing loss).

Self-management of hearing loss requires motivation and dedication, and, as a result, people become aware of their hearing difficulties (Henshaw et al., 2015). It could be that adherence and uptake of hearing aid technology could be higher with additional hearing care services (low-friction, easy to access, remote monitoring, training, and follow-up), which would also mitigate demands placed on the current rehabilitation centers (Lesica, 2018).

Extending the training duration or adding a maintenance phase could also enhance performance. A recent review by Chae & Bahng (2023) on the duration showed that the largest effect size was found for training periods longer than 600 minutes compared to periods shorter than 600 minutes. The brain might need more time for neural modifications to impact subjective experience. Long-term engagement could solidify benefits and improve generalization to untrained materials. Overall, participants adhered to the daily practice time requested, with some even willing to extend it. In the current study training time was at least 600 minutes (8 weeks, 15 min/day), while in our previous studies it was 1200 minutes (16 weeks, 15 min/day, Magits et al., 2023) and 900 minutes (12 weeks, 15 min/day, Van Wilderode et al. 2023). All three studies failed to show an improvement in subjective experience.

Regardless of the training gains, ALICE provides a service that can be broadly utilized, particularly in settings where access to traditional hearing healthcare is limited. Self-guided programs serve as a low-barrier supplement or alternative to traditional care. Monitoring performance through digits in noise and phoneme identification in quiet can be done remotely, allowing healthcare providers to follow up on progress without additional workload. Performance on the digits in noise task is highly correlated with sentence understanding in noise (van Wieringen et al., 2021) and can be done remotely, without requiring the patient to travel. In the current study, we presented the percentage correct scores for vowels and consonants. More detailed analyses of the confusion between phonemes are provided in the dashboard of the HCP. Information transmission analysis (Miller and Nicely, 1955) applied to the data of the four most recent stimulus-response matrices yields detailed insight into difficult-to-perceive speech features (van Wieringen et al. 2021). The HCP can track adherence to monitoring and training, generate alerts about clients with decreasing listening performance, and identify clients experiencing problems.

Regarding training, the dashboard provides information about the client’s performance on specific exercises and exercise types. The counseling module offers insights into the client’s perceived listening ability and satisfaction in daily life. This additional information helps healthcare providers (HCPs) deliver optimal care, enhance daily listening experiences, and improve the ability to manage challenging listening situations.

The ALICE app was proven safe during the multicenter clinical trial, with no data leakages and strong data privacy and security measures. No issues were reported regarding inaccurate sound levels. Despite a few initial bugs, the ALICE interface and the dashboard were user-friendly and easy to navigate. Participants could report issues as needed, and the app was regularly updated with the latest technology and security enhancements.

Although significant challenges remain in optimizing training programs, outcomes, and other factors, self-guided training programs offer greater personalization and interactivity. These programs promote self-management of listening problems and contribute to more equitable hearing care by increasing access to services. In the current study, hearing-impaired participants used hearing aids and/or cochlear implants. In the future, the efficacy of ALICE can also be evaluated in individuals without hearing interventions who have difficulty listening in noise. Additionally, streaming speech sounds directly to the ear via a hearing aid or cochlear implant may help improve hearing in the deaf ear, particularly in cases of single-sided deafness. Practicing solely with the deaf ear cannot be easily done in the clinic or free-field unless the good ear is blocked. ALICE is available in the Dutch and French languages.

## Conclusion

Our clinical trial shows that the self-guided ALICE training program enhances DiN and phoneme perception, and is effective at improving the auditory system’s ability to parse untrained speech in noise. This enhancement in speech in noise performance is specific to the training group, as the control group did not improve. Participants were compliant during the 8-week training period. The results of the clinical trials imply that ALICE can be used as a scalable, accessible, and safe hearing care intervention.

## Data Availability

All data produced in the present work are contained in the manuscript

## Acknowledgments

We sincerely thank the hearing centers Aerts, Amplifon, Audika, Jolien Desmet, Charlotte Marinus, and the university hospital Leuven for their time and effort to recruit and test participants. We also thank Dr Andrea Bussé for her assistance with preparing the clinical trial and training the investigators. Stefanie Krijger, dr Tilde Van Hirtum, dr Laura Leyssens are thanked for their assistance with the regulatory work in the first phase of the project. This work was supported by a C3-grant project from the KU Leuven (C3/21/046) and by the FWO-TBM grant no T000823N (CLINIC, PI van Wieringen)

## Author Contributions

AvW and MVW contributed to the study design and performed the data analyses. LDR programmed the ALICE app. AvW, MVW, LDR, TF, and JW provided critical revision and feedback. The study protocol is registered on ClinicalTrials.gov ID: NCT05329922.

